# The burden of the postictal state in epilepsy: a prospective, single-centre observational cohort study

**DOI:** 10.64898/2026.03.20.26348929

**Authors:** Ionu-Flavius Bratu, Agnès Trébuchon, Fabrice Bartolomei

**Affiliations:** Department of Epileptology and Cerebral Rhythmology, Timone Hospital, 13005, Marseille, France; Systems Neuroscience Institute, Aix-Marseille University, French National Institute of Health and Medical Research (UMR 1106), 13005, Marseille, France

**Keywords:** postictal state, postictal recovery, scale, duration, burden

## Abstract

**Objective:** The postictal state is a major yet underrecognised component of epilepsy burden. We aimed to develop a structured patient-reported instrument to quantify postictal recovery, characterise its multidimensional burden and identify demographic, clinical, psychiatric and treatment-related factors associated with postictal severity and duration.

**Methods:** We conducted a prospective, single-centre observational cohort study (Timone Hospital, Marseille, February 2025 - March 2026). Consecutive patients aged ≥15 years admitted for scalp or stereo-EEG video-monitoring were included. Patients completed the Postictal Recovery Scale (PRS), an 11-domain questionnaire assessing fatigue, mood, sensory, motor, language, orientation, time perception and postictal amnesia. Items were rated from 0 (severe impairment) to 3 (no symptoms), yielding a total score of 0-33. Internal consistency was assessed using Cronbach’s alpha. Associations between PRS scores, subjective postictal duration and covariates were analysed using group comparisons, correlations and regression models.

**Results:** Of 107 enrolled patients, 96 were included. PRS showed good internal consistency (Cronbach’s α = 0.79). 96% of patients reported experiencing postictal symptoms, with fatigue (80%) and postictal amnesia (79%) being the most frequent and severe manifestations. Recovery exceeded one hour in 21% of patients. Greater postictal impairment was associated with higher interictal anxiety (Spearman ρ = −0.32, p = 0.0018) and depressive symptoms (Spearman ρ = −0.40, p = 0.0001), whereas demographic, epilepsy-related and treatment variables showed no significant associations. Altered postictal time perception was reported by 40% of patients and was associated with disorientation, but not psychiatric symptoms. Subjective postictal duration was longer than subjective ictal duration (Wilcoxon test, p < 0.0001).

**Significance:** The postictal state is a frequent and multidimensional patient-reported experience. Greater postictal severity, particularly concerning anxiety and depression, is associated with interictal psychiatric comorbidity, while altered temporal experience emerges as a distinct dimension of postictal dysfunction. These findings support integrating postictal measures into clinical practice and trials.

## 1. Introduction

Although seizures represent the defining clinical manifestation of epilepsy, the overall burden of the disorder extends beyond the ictal event itself and includes consequences occurring before, during and after seizures.^1^ Among these peri-ictal phases, the postictal state is the transient abnormal cerebral condition that begins after seizure termination and persists until return to interictal baseline.^2^ It may include fatigue, cognitive impairment, language disturbances, sensorimotor deficits and neuropsychiatric symptoms^3^, reflecting the disruption of large-scale brain networks following seizures.^4^ Importantly, postictal consequences may extend well beyond the seizure itself.^5–7^ While seizures last seconds to minutes, postictal symptoms may persist for hours or longer.^8,9^ This functional and temporal dissociation suggests that the cumulative burden of epilepsy may arise less from the ictal event than from its aftermath.^10^ Despite this, the postictal phase remains understudied and is rarely assessed systematically in routine clinical practice.^11^ One major reason is the lack of structured tools capable of capturing its multidimensional nature.^12^ Postictal-specific instruments remain rare^4^ and most available seizure-related scales focus on ictal semiology and severity, with only limited consideration of postictal manifestations.^13–15^ As a result, postictal symptoms are often described in relation to isolated phenomena rather than being quantified across domains and the factors associated with postictal severity and duration remain poorly characterised.^8,9^

Temporal processing underlies behaviour and decision-making, both important determinants of quality of life. Evidence suggests that explicit timing may be impaired in epilepsy, whereas implicit timing mechanisms are relatively preserved.^16^ Moreover, temporal orientation is a core component of postictal confusion and is typically among the last functions to recover after a seizure.^17^ Together, these observations suggest that alterations in temporal experience may contribute to the subjective burden of epilepsy, especially during the postictal state.

Patient-reported outcomes are increasingly recognised as essential components of epilepsy care and research, as they capture dimensions of disease burden not fully reflected by conventional clinical metrics alone.^18^ This is particularly relevant for the postictal state, whose impact on daily functioning and quality of life may be substantial yet remains insufficiently quantified.^11,19^ The main aim of the present study was to develop a structured patient-reported instrument to quantify postictal recovery, the Postictal Recovery Scale (PRS). Secondary objectives were to characterise the multidimensional burden of the postictal state and to investigate demographic, clinical, epilepsy-related, comorbidity-related and treatment-related factors associated with postictal severity and recovery duration. Finally, we explored whether peri-ictal alterations in time perception may represent an under-recognised contributor to the overall ictal–postictal burden.

## 2. Methods

### 2.1 Study design and setting

This prospective observational study was conducted in the Department of Epileptology and Cerebral Rhythmology at Timone Hospital, Marseille, France, between February 2025 and March 2026, in the context of routine admissions to the Epilepsy Monitoring Unit (EMU). All participants provided written informed consent prior to inclusion. The study was conducted in accordance with the Declaration of Helsinki and was approved by the local ethics committee of Assistance Publique–Hôpitaux de Marseille (AP-HM) (PADS25-96).

### 2.2 Participants

All consecutive patients hospitalised in the EMU during the study period were considered for inclusion, regardless of video-monitoring modality (scalp or stereo-EEG). Patients were eligible if they were aged ≥15 years and had a baseline neurocognitive status compatible with reliable questionnaire completion. Patients were excluded from the final analysis if the events recorded during monitoring were classified as functional seizures only, if epileptic and functional seizures coexisted, but could not be reliably distinguished in symptom reporting, or if only other transient non-epileptic events (e.g., syncope) were recorded. Exclusions were applied after monitoring completion, once the electroclinical diagnosis had been established.

### 2.3 Postictal recovery questionnaire

Participants completed a structured self-administered questionnaire developed for this study to assess subjective postictal recovery following epileptic seizures. The questionnaire – Postictal Recovery Scale **(PRS, Suppl.mat. – Table 1)** – was developed based on current literature on postictal manifestations^3,8^ and the clinical experience of the authors. PRS assesses fatigue, emotional symptoms, sensory disturbances, motor or coordination deficits, spoken language comprehension and production, temporo-spatial orientation, perception of the passage of time and postictal amnesia. Emotional symptoms were explored through separate items addressing sadness/apathy, anxiety and elevated/euphoric mood. Each item was rated from 0 (severe impairment) to 3 (no impairment), yielding a total score of 0–33. Open fields allowed patients to specify symptoms such as lateralisation and localisation of motor/sensory deficits or additional postictal phenomena not otherwise captured (e.g., headache). Patients also estimated the typical time needed to return to baseline using predefined categories (0–5, 5–10, 10–15, 15–30, 30–60 minutes, or >1 hour), with optional free-text entries for greater precision.

Patients were instructed to complete the questionnaire with reference to the seizure type and severity they considered most representative of their usual epileptic events. PRS was administered following EMU admission once both patient report and clinical examination confirmed absence of ongoing postictal signs and symptoms. Information on habitual seizure duration was also collected and, when available, objectively measured from video-EEG or SEEG recordings. Seizure onset was defined as the earliest clinical or electrographic onset and seizure end as electrographic offset; when several habitual seizures were recorded, the mean duration was used. In a subgroup of patients, PRS was also administered shortly after seizures recorded during monitoring, typically within 30 minutes of seizure termination.

### 2.4 Statistical analysis

Statistical analyses were performed using GraphPad Prism version 11 and Jamovi version 2.7.23. Continuous variables are reported as mean ± standard deviation and range and categorical variables as counts and percentages. Normality was assessed using the Shapiro–Wilk test. Group comparisons were performed using Student’s t-tests or one-way ANOVA for normally distributed variables and Mann–Whitney U or Kruskal–Wallis tests for non-parametric data, with Tukey or Dunn post-hoc tests where appropriate. Associations between continuous or ordinal variables were assessed using Spearman rank correlations. PRS internal consistency was evaluated using Cronbach’s alpha coefficient, with item–rest correlations and alpha-if-item-deleted values also examined. Subjective postictal recovery duration was coded as an ordinal variable from 1 to 6. To explore the relationship between treatment-related factors and postictal severity, multivariable linear regression models were constructed with PRS total score as the dependent variable and the number of anti-seizure medications (ASM) and their pharmacological classes as predictors.^20^ Predictor variables were entered simultaneously into the model. Ordinal logistic regression models were used for PRS subdomain scores and subjective postictal duration. ASMs were classified by their principal mechanism of action.^20^ **(Suppl.mat. – Table 2).** All tests were two-tailed, with significance set at p < 0.05; Bonferroni correction was applied when appropriate.

## 3. Results

### 3.1 Study population

A total of 107 patients were prospectively enrolled. After application of the predefined exclusion criteria, 11 patients were excluded: seven with coexisting focal drug-resistant epilepsy and functional (dissociative) seizures, two with only functional (dissociative) seizures recorded during monitoring, one with syncopal events only and one patient with Lennox-Gastaut syndrome, precluding a representative postictal assessment. Therefore, the final cohort comprised 96 patients.

### 3.2 Demographic and clinical characteristics

We analysed the data from 51 women (53%) and 45 men (47%), with a mean age of 35.9 ± 12.2 years (range: 16–87). The mean age at epilepsy onset was 19.1 ± 14.4 years (range: 0.1–76), corresponding to a mean epilepsy duration of 16.8 ± 11.9 years (range: 1–56). Patients experienced a mean seizure frequency of 14.1 ± 25.5 seizures per month (range: 0–135). Focal to bilateral tonic–clonic seizures were present in 25 patients (26%), representing 37.2 ± 41.6% of overall seizure burden (range: 0.4–100%). Patients were treated with a mean of 2.6 ± 1 ASMs (range: 0–5). The epileptogenic zone network (EZN) was unilateral in most patients (86/96, 90%), with left-sided EZN in 55 (57%) and right-sided in 31 (33%), while 10 (10%) had bilateral EZN. Most EZNs were unilobar, predominantly involving the temporal lobe (60 patients, 63%), whereas multilobar EZNs were present in 21 patients (22%). The most frequently prescribed ASMs were lacosamide (44%), cenobamate (35%), lamotrigine (25%), carbamazepine (22%) and levetiracetam (21%) **(Suppl.mat. – Table 2)**. Vagus nerve stimulation was present in 9 patients (9%), while 6 patients (6%) had undergone prior resective epilepsy surgery. The demographic and clinical characteristics are summarised in **Table 1**.

**Table 1.** Demographic and clinical characteristics of the study cohort (n = 96)

| Variable | Value |
| --- | --- |
| **Sex** |  |
| Women | 51 (53%) |
| Men | 45 (47%) |
| **Age (years)** | 35.9 ± 12.2 (16–87) |
| **Age at epilepsy onset (years)** | 19.1 ± 14.4 (0.1–76) |
| **Epilepsy duration (years)** | 16.8 ± 11.9 (1–56) |
| **Seizure frequency (per month)** | 14.1 ± 25.5 (0–135) |
| **Patients with focal-to-bilateral tonic–clonic (FTBTC) seizures** | 25 (26%) |
| Proportion of FTBTC seizures within monthly seizure frequency | 37.2 ± 41.6% (0.4–100) |
| **Number of antiseizure medications** | 2.6 ± 1.0 (0–5) |
| **Epileptogenic zone network** |  |
| Unilateral | 86 (90%) |
| Left | 55 (57%) |
| Right | 31 (33%) |
| Bilateral | 10 (10%) |
| **Lobar localisation of the epileptogenic zone network** |  |
| Temporal | 60 (63%) |
| Frontal | 9 (9%) |
| Parietal | 3 (3%) |
| Occipital | 2 (2%) |
| Insular | 1 (1%) |
| Multilobar | 21 (22%) |
| **Anti-seizure medication** |  |
| Sodium-channel blocker | 79 (82%) |
| SV2A ligand | 39 (41%) |
| GABAergic | 25 (26%) |
| Mixed or other mechanisms | 55 (75%) |
| **Previous epilepsy surgery** |  |
| Yes | 6 (6%) |
| No | 90 (94%) |
| **Ongoing vagus nerve stimulation** |  |
| Yes | 9 (9%) |
| No | 87 (91%) |

### 3.3 Postictal Recovery Scale – quantitative results

The internal consistency of PRS was good (Cronbach’s α = 0.79). Item–rest correlations ranged from 0.22 to 0.60 and removal of individual items did not improve internal consistency.

The mean PRS global score was 23.3 ± 6.1 (range: 8–33), indicating a substantial postictal burden across the cohort, with 92 patients (96%) reporting at least one postictal disturbance (PRS total score <33). The lowest mean scores were observed for fatigue and postictal amnesia, which were also the most frequent symptoms (80% and 79%, respectively). Negative emotional symptoms were common (depressive/apathy: 61%; anxiety: 68%), whereas euphoria was uncommon (15%). Cognitive disturbances were also frequent, including temporo-spatial disorientation (46%), language production deficits (43%) and comprehension deficits (38%). Moreover, motor and sensory disturbances were reported by 45% and 35% of patients, respectively. Detailed PRS domain scores and frequencies are summarised in **Table 2**.

**Table 2.** Postictal Recovery Scale (PRS) scores and subjective postictal duration (n = 96)

| Variable | Mean ± SD (range) | n (%) |
| --- | --- | --- |
| **PRS total score** | 23.3 ± 6.1 (8–33) | 92 (96%)* |
| **PRS domains** |  |  |
| Fatigue | 1.7 ± 0.9 (0–3) | 77 (80%) |
| Depression / apathy | 1.8 ± 1.1 (0–3) | 59 (61%) |
| Anxiety | 1.9 ± 1.0 (0–3) | 65 (68%) |
| Euphoria | 2.8 ± 0.6 (0–3) | 14 (15%) |
| Sensory deficit | 2.5 ± 0.7 (1–3) | 34 (35%) |
| Motor deficit | 2.3 ± 0.9 (0–3) | 43 (45%) |
| Comprehension deficit | 2.4 ± 0.9 (0–3) | 36 (38%) |
| Production deficit | 2.3 ± 1.0 (0–3) | 41 (43%) |
| Temporal-spatial orientation | 2.1 ± 1.2 (0–3) | 44 (46%) |
| Passage of time judgement | 2.3 ± 1.1 (0–3) | 38 (40%) |
| Postictal amnesia | 1.2 ± 1.2 (0–3) | 76 (79%) |
| **Subjective postictal recovery duration** |  |  |
| 0–5 minutes | - | 29 (30%) |
| 5–10 minutes | - | 14 (15%) |
| 10–15 minutes | - | 8 (8%) |
| 15–30 minutes | - | 14 (15%) |
| 30–60 minutes | - | 9 (9%) |
| >1 hour | - | 20 (21%) |
| Unable to estimate | - | 2 (2%) |

PRS items were rated on a four-point ordinal scale ranging from 0 to 3, where 0 indicates severe impairment and 3 indicates normal function.
\*The PRS total score ranges from 0 to 33; therefore, scores <33 indicate the presence of at least one postictal symptom.

To explore associations between postictal symptom domains, pairwise Spearman rank correlations between the PRS domains were calculated (**Fig. 1**). After Bonferroni correction, several associations remained significant. The strongest was between speech comprehension and production deficits (ρ = 0.68). Language impairments were also associated with temporo-spatial disorientation (ρ = 0.47 and ρ = 0.42) and time perception disturbances (ρ = 0.45 and ρ = 0.41). Motor deficits correlated with fatigue (ρ = 0.44), depression (ρ = 0.41), speech production deficits (ρ = 0.44), temporo-spatial disturbances (ρ = 0.37) and time perception alterations (ρ = 0.39). Postictal depression and anxiety were associated (ρ = 0.50), whereas postictal amnesia showed no significant correlations after correction.

**Figure 1.**
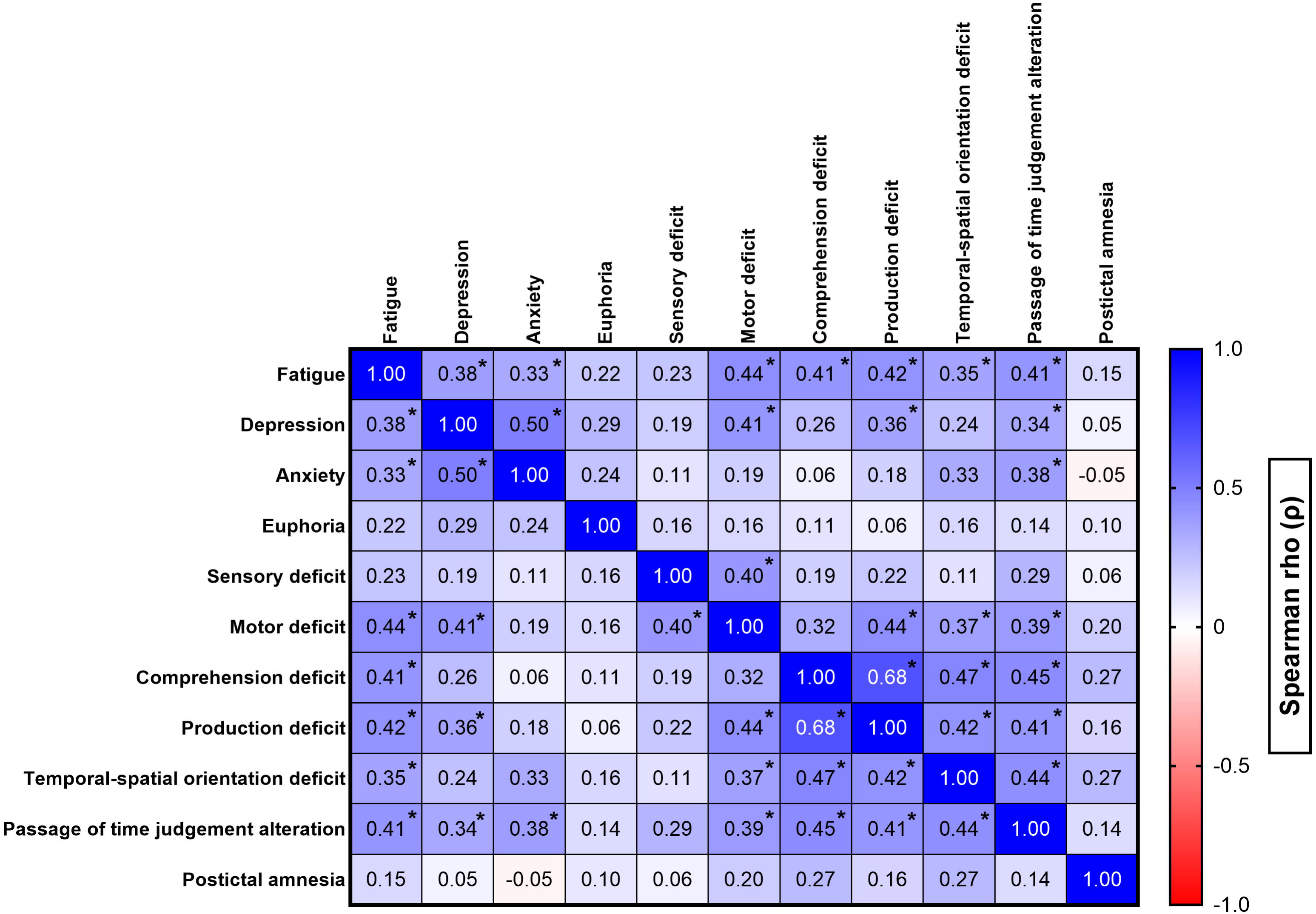
Correlation heatmap of postictal symptom domains. Pairwise Spearman rank correlation coefficients (ρ) between the eleven domains of the Postictal Recovery Scale are displayed. Blue colours indicate positive correlations and red colours indicate negative correlations, with colour intensity reflecting the strength of the association. Asterisks denote correlations that remained significant after Bonferroni correction for multiple comparisons.

Patient-reported postictal recovery time varied substantially across the cohort. The most frequent interval was 0–5 minutes (30%), but 21% of patients reported postictal states lasting more than one hour (**Table 2**). A subgroup of 33 patients provided free-text estimates of the duration of specific postictal symptoms or their overall recovery. Among these, 45% reported very brief symptoms ranging from immediate recovery to several minutes, whereas 27% described symptoms persisting for persisting from one hour to several hours and another 27% reported disturbances lasting one day or longer, most often characterised by prolonged fatigue or cognitive slowing. One patient reported recovery lasting up to six weeks, although the symptoms underlying this prolonged course were not specified.

### 3.4 Postictal Recovery Scale – qualitative responses

Open-text fields provided additional insight beyond structured PRS domains. The most frequent additional symptoms were headache (11 patients; 11.5%) and blurred vision (8 patients; 8.3%). Other phenomena were rare and included depersonalisation-like experiences, a non-specific feeling of being “off”, postictal wandering, bitter taste, metallic smell, tinnitus, transient hearing impairment, short-term memory difficulties and sleep phase shift, each reported by a single patient.

### 3.5 Postictal severity and duration – demographic and epilepsy-related factors

Neither PRS scores (total or subdomain) nor subjective postictal duration differed across sex, EZN hemispheric distribution (unihemispheric vs bihemispheric), EZN laterality (left vs right), or lobar organisation (unilobar vs multilobar) (Mann–Whitney U tests, p > 0.05). Before Bonferroni correction, trends included sex differences in postictal anxiety (Mann–Whitney U test, p = 0.03) and greater comprehension deficits in unilobar temporal versus extratemporal EZN (Mann–Whitney U test, p = 0.0185). Furthermore, PRS scores and subjective postictal duration were not associated with age, age at epilepsy onset, epilepsy duration or monthly seizure frequency (Spearman correlations, p > 0.05). Similar results were observed for focal-to-bilateral tonic–clonic seizure frequency (PRS scores - total or subdomain: n = 25; subjective postictal duration: n = 24; Spearman correlations, p > 0.05).

### 3.6 Postictal severity and duration – treatment-related factors

Patients with prior epilepsy surgery had higher PRS total scores (Mann–Whitney U test, p = 0.0059), with similar trends for fatigue (p = 0.0360), motor deficit (p = 0.0426) and postictal duration (p = 0.0075). However, these differences did not remain significant after Bonferroni correction. PRS scores (total or subdomain) and postictal duration did not differ between patients with and without vagus nerve stimulation therapy (Mann–Whitney U tests, p > 0.05). Moreover, multivariable linear regression analyses did not reveal associations between PRS total score and either the number of antiseizure medications or ASM pharmacological classes (p > 0.05). Similarly, ordinal logistic regression analyses did not identify associations between these treatment variables and individual PRS subdomains or postictal duration (p > 0.05).

### 3.7 Postictal severity and duration – psychiatric correlates

Psychiatric symptoms were assessed using the Generalized Anxiety Disorder-7 (GAD-7) and the Neurological Disorders Depression Inventory for Epilepsy (NDDI-E). Psychiatric screening revealed a mean GAD-7 score of 8.7 ± 5.6 (range: 0–21) and a mean NDDI-E score of 12.0 ± 4.4 (range: 6–23). PRS total scores were associated with interictal anxiety levels and depressive symptoms. Higher anxiety levels were associated with greater postictal impairment, as reflected by a significant correlation between PRS and GAD-7 scores (Spearman ρ = −0.32, p = 0.0018, n = 91). PRS scores also differed across GAD-7 anxiety severity categories (one-way ANOVA, F(3,87) = 3.12, p = 0.030), with Tukey post-hoc comparisons indicating lower PRS values in patients with severe anxiety compared with those with minimal anxiety (p = 0.016) (**Fig. 2A**). Similarly, NDDI-E scores were correlated with PRS values (Spearman ρ = −0.40, p = 0.0001, n = 91). Patients scoring above the NDDI-E screening threshold for depression (NDDI-E ≥13) exhibited significantly lower PRS scores than those below the threshold (Mann–Whitney U test, p = 0.0003).

**Figure 2.**
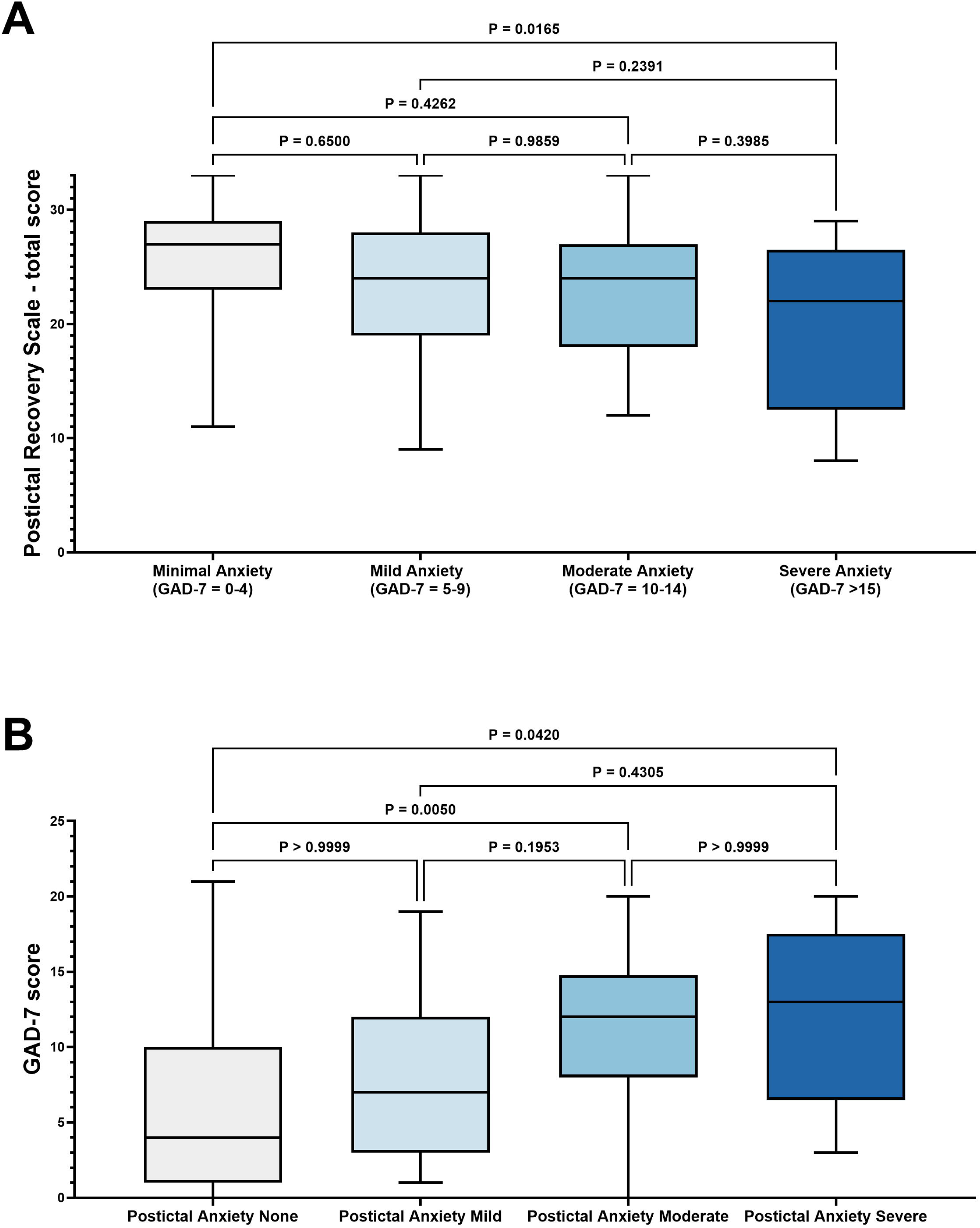
Relationship between interictal anxiety severity and postictal impairment. **(A)** Distribution of Postictal Recovery Scale (PRS) total scores across categories of interictal anxiety severity as measured by the Generalized Anxiety Disorder-7 (GAD-7) questionnaire (minimal, mild, moderate and severe anxiety). Lower PRS scores indicate greater postictal impairment. **(B)** Distribution of GAD-7 scores according to the severity of postictal anxiety reported in the PRS (none, mild, moderate, severe). Box plots show the median and interquartile range, with whiskers representing the range of observed values. Statistical comparisons were performed using one-way ANOVA with Tukey post-hoc tests in panel A and Kruskal–Wallis tests with Dunn post-hoc comparisons in panel B.

**Figure 3.**
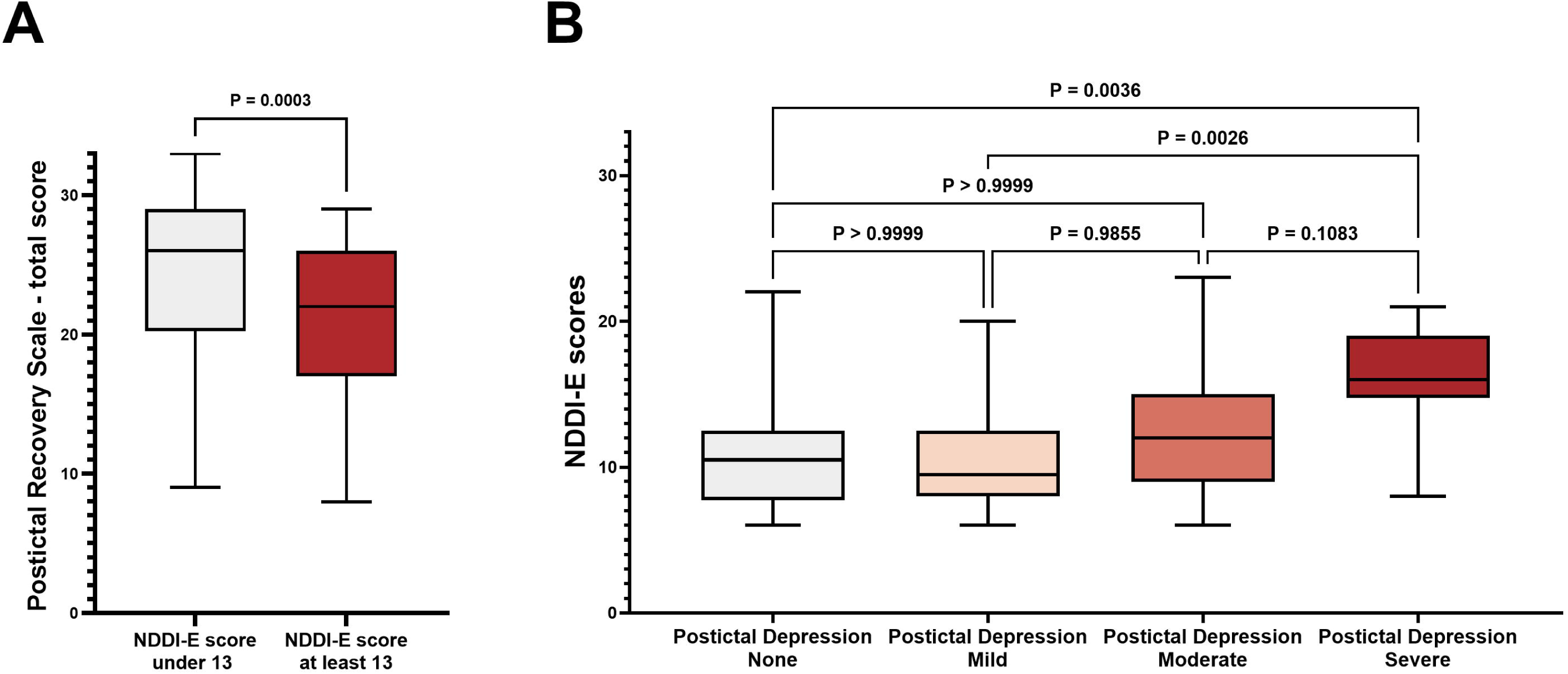
Relationship between interictal depressive symptoms and postictal impairment. **(A)** Distribution of Postictal Recovery Scale (PRS) total scores according to depressive symptom screening using the Neurological Disorders Depression Inventory for Epilepsy (NDDI-E), dichotomised at the standard screening threshold (NDDI-E < 13 vs ≥ 13). **(B)** Distribution of NDDI-E scores according to the severity of postictal depressive symptoms reported in the PRS (none, mild, moderate, severe). Boxes represent the interquartile range (25th–75th percentile), the horizontal line indicates the median and whiskers represent the minimum and maximum values. Individual points correspond to individual patients. Statistical comparisons were performed using a Mann–Whitney U test in panel A and a Kruskal–Wallis test with Dunn post-hoc comparisons in panel B. Lower PRS scores indicate greater postictal impairment.

Postictal anxiety and depressive severity were associated with interictal anxiety and depressive symptoms. Increasing postictal anxiety severity correlated with higher GAD-7 scores (Spearman ρ = −0.40, p = 0.0001, n = 91) and GAD-7 scores differed across postictal anxiety severity groups (Kruskal–Wallis test, p = 0.0023). Post-hoc Dunn comparisons showed significantly higher scores in patients with moderate (p = 0.005) and severe (p = 0.042) postictal anxiety compared with those without postictal anxiety (**Fig. 2B**). Postictal depressive severity correlated with NDDI-E scores (Spearman ρ = −0.32, p = 0.0021, n = 91) and NDDI-E scores differed across postictal depression severity groups (Kruskal–Wallis test, p = 0.0019). Dunn post-hoc tests revealed significantly higher scores in patients with severe postictal depression compared with those with no or mild postictal depressive symptoms (p = 0.0036 and p = 0.0026).

Subjective postictal recovery duration was not correlated with either GAD-7 or NDDI-E scores (Spearman correlations, p > 0.05, n = 80).

### 3.8 Postictal severity and duration – chronic and acute perception

In a subgroup of 10 patients assessed within 30 minutes of seizure termination, acute PRS scores (total or subdomain) and subjective postictal recovery duration did not differ from retrospectively reported values (paired Wilcoxon signed-rank tests, all p > 0.05).

### 3.9 Ictal-postictal temporal perspective

Alterations in the subjective perception of the passage of time during the postictal period were reported by 38 patients (40%). The mean PRS score for the passage-of-time judgement domain was 2.3 ± 1.1 (range: 0–3), indicating that while many patients experienced some degree of temporal distortion following seizures, these disturbances were generally mild to moderate. Disturbances in temporal perception were moderately associated with postictal temporo-spatial orientation deficits (Spearman ρ = 0.44, p = 0.006). In contrast, the passage-of-time perception domain was not significantly correlated with postictal anxiety or postictal depressive symptoms (Spearman correlations, p > 0.05).

To further explore seizure-related temporal perception, subjective estimates of seizure duration were compared with objectively measured seizure durations from video-EEG recordings (scalp EEG or SEEG). Self-reported seizure duration averaged 2.4 ± 3.9 minutes (range: 0.1–30), whereas objectively measured seizure duration averaged 1.4 ± 1.0 minutes (range: 0.1–5). Subjective seizure duration differed significantly from objectively measured seizure duration (paired Wilcoxon signed-rank test, p = 0.011, n = 48).

A subgroup of patients provided free-text estimates for both subjective seizure duration and postictal recovery duration (n = 30). Free-text estimates of postictal duration were substantially longer than ictal durations (Wilcoxon matched-pairs signed-rank test, p < 0.0001). However, ictal and postictal durations were not significantly correlated (Spearman correlation, p > 0.05). This lack of association persisted when ordinally transformed postictal duration intervals were correlated with subjective ictal durations in the larger cohort (Spearman correlation, p > 0.05, n = 74) (**Fig. 4**).

**Figure 4.**
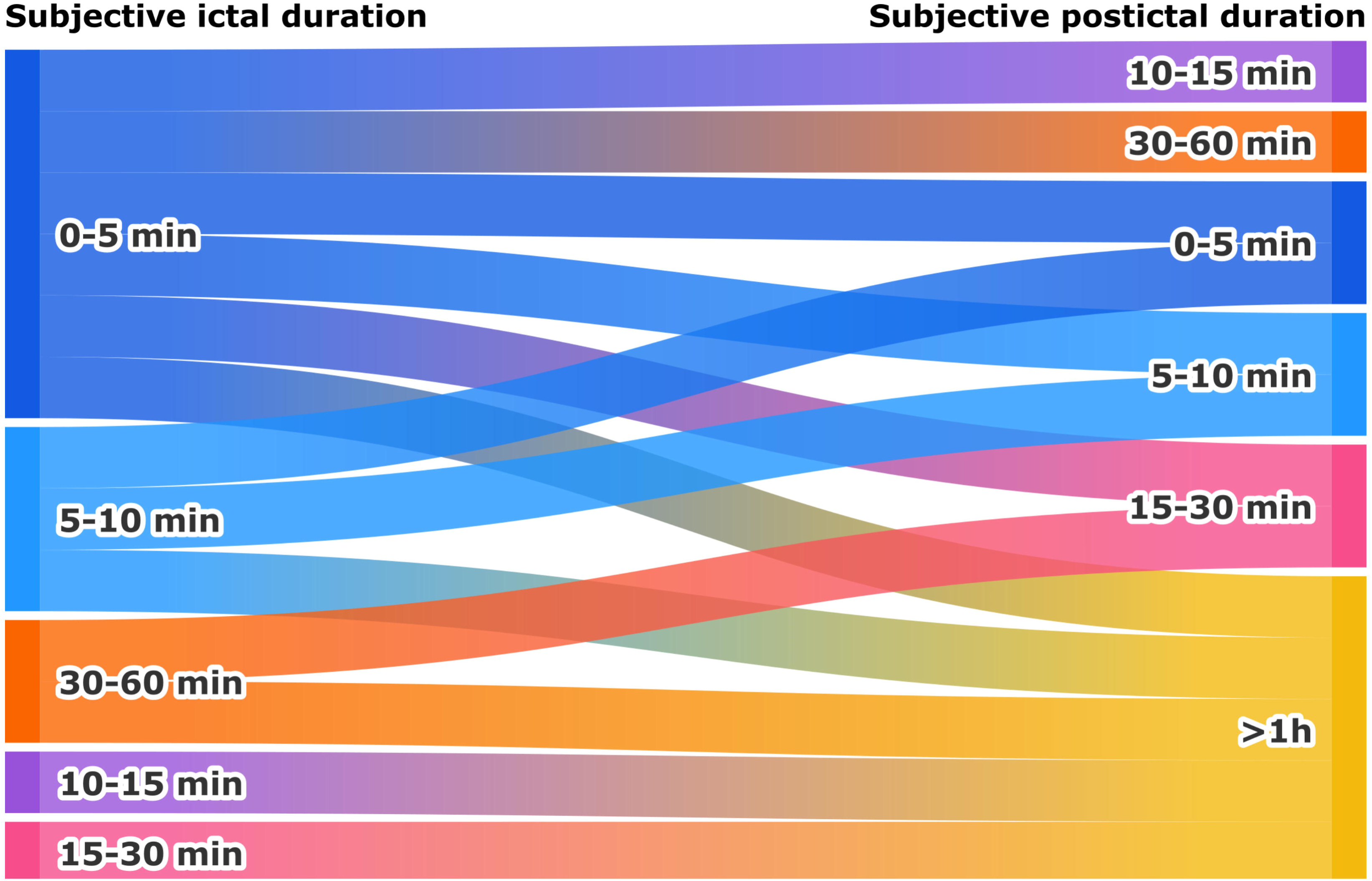
Relationship between subjective ictal duration and subjective postictal recovery duration. Sankey diagram illustrating the distribution of patients across combinations of self-reported seizure duration categories and subjective postictal recovery duration categories. Ribbon width is proportional to the number of patients reporting each combination of categories. The heterogeneous distribution of ribbons highlights the absence of a consistent relationship between perceived seizure duration and postictal recovery time.

## 4. Discussion

To our knowledge, this is the first study to develop and apply a structured patient-reported multidomain instrument specifically designed to quantify postictal recovery in epilepsy. Using the Postictal Recovery Scale in a prospective EMU cohort, we found that postictal symptoms were almost universal, multidimensional and often prolonged. Fatigue and postictal amnesia emerged as the most frequent and severe manifestations, while affective, language, orientation and sensorimotor disturbances were also common. Greater postictal burden was associated with higher interictal anxiety and depressive symptom scores, whereas most demographic, epilepsy-related and treatment-related variables were not. Finally, altered temporal experience emerged as a relevant component of the postictal state and perceived recovery exceeded seizure duration.

Unlike existing seizure severity instruments, which only indirectly capture postictal and experiential aspects of seizures^13,21^, PRS was designed specifically to quantify patient-reported recovery across multiple domains. In this cohort, it showed good internal consistency, supporting its use as a composite measure of postictal dysfunction. Nearly all patients reported at least one postictal symptom, with fatigue and postictal amnesia standing out as central features of postictal burden. Fatigue may reflect the combined impact^22,23^ of seizure-related physiological stress, postictal sleepiness and mood-related factors, whereas postictal amnesia may result from dysfunction within memory-related networks or incomplete awareness during the peri-ictal period.^4,7,24,25^ More broadly, PRS domain correlations indicate that postictal symptoms co-occur as interrelated dysfunctions. This aligns with prior findings showing that functional recovery parallels recovery within overlapping networks, supporting network-level reinstatement during postictal recovery.^4,26^

Qualitative responses identified additional symptoms (e.g., headache, sensory disturbances, transient cognitive or perceptual changes and sleep-phase shift), underscoring the phenomenological richness of the postictal state and supporting the inclusion of open-text fields alongside structured instruments. In ambulatory settings, the postictal period may be the most accessible phase for direct clinical examination beyond the clinical history and may provide information for EZN lateralisation and localisation^4^, as well as contribute to the differential diagnosis of paroxysmal events.^27^ Moreover, certain symptoms carry functional and safety implications: postictal wandering or agitation may expose patients to self-endangerment, while residual symptoms, including sleep-phase shift, may delay return to daily activities, including work and school.^5,11,28,29^

One notable finding was the absence of robust associations between postictal burden and most conventional demographic and epilepsy-related variables. Sex, age, age at epilepsy onset, epilepsy duration, seizure frequency, EZN laterality, EZN hemispheric distribution and lobar organisation were not significantly associated with PRS scores or subjective postictal duration. Although an uncorrected trend suggested greater postictal speech comprehension impairment in unilobar temporal compared with unilobar extratemporal epilepsy, this did not survive correction for multiple comparisons. These findings only partly accord with our previous SEEG study of objective postictal recovery, in which recovery was associated with lobar organisation, seizure duration, age, age at epilepsy onset, bilateral EZN organisation and histopathology, whereas EZN laterality and epilepsy duration were not significant predictors.^9^ This discrepancy may reflect differences between objective electrophysiological recovery and the lived subjective burden captured by PRS. Likewise, neither the number of ASMs, broad pharmacological classes nor ongoing vagus nerve stimulation was associated with postictal burden and the apparent association with prior epilepsy surgery did not survive correction. These findings should be interpreted cautiously because patients receiving neuromodulation or prior surgery were few and ASM combinations were heterogeneous, limiting statistical power and comparability with the existing literature.^30,31^

By contrast, the postictal burden showed consistent relationships with interictal psychiatric symptoms. Higher GAD-7 and NDDI-E scores were associated with lower PRS total scores and greater postictal anxiety and depressive severity were themselves linked to higher interictal anxiety and depressive symptom scores. These findings suggest that the subjective burden of the postictal state is closely intertwined with psychiatric vulnerability. Several mechanisms may contribute to this relationship. First, interictal anxiety and depression may amplify the salience, distress, or memorability of postictal symptoms.^32^ Second, limbic and paralimbic network dysfunction may underpin a bidirectional relationship between psychiatric comorbidity and peri-ictal burden, whereby pre-existing affective vulnerability amplifies seizure-related symptoms and recurrent seizures may in turn reinforce emotional-network dysfunction.^33,34^ Third, mood symptoms, fatigue, poor sleep and reduced resilience may interact to worsen the perceived aftermath of seizures.^35^ Interestingly, psychiatric symptoms were associated with PRS severity, but not with subjective postictal duration, suggesting they may relate more closely to the affective-experiential burden of the postictal state than to its perceived temporal extent. Our results should, however, be considered in light of the predominance of EZNs involving the temporal lobe in our cohort, as temporal lobe epilepsy is known to carry a particularly high burden of psychiatric comorbidity. ^36,37^

Although exploratory, the comparison between acute and retrospective PRS assessments is also of interest. In the small subgroup assessed shortly after seizure termination, acute PRS scores and subjective recovery duration did not differ significantly from retrospectively reported habitual values. This suggests that, in at least some patients, interictal reports may provide a reasonably stable summary of habitual postictal experience, which may be useful when immediate postictal assessment is not feasible.

Finally, the temporal perception findings add an original dimension to the present study. Most epilepsy literature on time perception concerns the interictal and ictal states.^16,38–40^ In our study, altered judgement of the passage of time was reported by 40% of patients and was associated with temporo-spatial disorientation, supporting the idea that disturbed time experience forms part of the broader postictal cognitive phenotype. Interestingly, the passage-of-time perception domain was not associated with postictal anxiety or depressive symptoms, suggesting that altered temporal experience may not be driven primarily by postictal affective factors. In addition, subjective seizure duration was significantly longer than objectively measured seizure duration, consistent with previous work showing that patient-reported spell duration may differ from recorded duration.^41^ However, subjective ictal duration was not correlated with subjective postictal recovery duration. This dissociation suggests that perceived seizure duration and perceived recovery duration are not interchangeable expressions of the same temporal distortion, but likely reflect distinct experiential constructs shaped by awareness, memory discontinuity, disorientation, affective state and variable recovery of large-scale brain networks. Subjective temporal estimates should therefore be considered part of the lived burden of epilepsy rather than simple proxies for objective duration, particularly as, in our study, subjective postictal duration outlasted subjective seizure duration, perhaps paralleling observations that postictal electrical disturbances outlast ictal ones and track recovery.^9,42^

## 5. Limitations

Several limitations should be acknowledged. First, this was a single-centre study conducted in a tertiary referral centre, likely enriched for drug-resistant focal epilepsies, which may limit generalisability. Second, although the PRS showed good internal consistency, it remains an exploratory instrument and has not undergone full psychometric validation, including factor analysis, test-retest reliability, responsiveness analysis or external validation in an independent cohort. Third, patients were asked to report their habitual postictal experience rather than complete separate questionnaires for each seizure type, which may introduce heterogeneity in patients with more than one relevant seizure phenotype. Moreover, they were instructed to answer with reference to the seizure severity they considered most representative of their habitual seizures. Fourth, patient-reported measures are inherently vulnerable to recall bias, impaired awareness and retrospective reconstruction.

## 6. Conclusion and perspectives

Beyond its descriptive value, the PRS may also have practical implications for both clinical care and research. In routine practice, PRS could help clinicians incorporate recovery burden into therapeutic decision-making. In clinical and therapeutic trials, PRS may provide a useful framework for capturing postictal morbidity as a patient-centred outcome measure. Although the present version was designed as a self-reported instrument, the same conceptual framework could also be adapted for proxy reporting by caregivers or healthcare professionals in situations where reliable self-report is limited, such as in children or patients with cognitive impairment. Future studies should assess whether patient- and caregiver-reported postictal measures converge and whether adapted PRS versions retain validity across epilepsy populations. Multicentre validation across centres with varying expertise will be required to establish generalisability, reproducibility and clinical applicability.

## Author contributions (CRediT)

**I-F.B.**: Conceptualization, Data curation, Formal analysis, Investigation, Methodology, Project administration, Software, Validation, Visualization, Writing – original draft, Writing – review and editing; **A.T.**: Conceptualization, Data curation, Funding acquisition, Investigation, Methodology, Project administration, Resources, Supervision, Validation, Writing – original draft, Writing – review and editing; **F.B.**: Conceptualization, Data curation, Funding acquisition, Investigation, Methodology, Project administration, Resources, Supervision, Validation, Writing – original draft, Writing – review and editing.

## Acknowledgements

We thank Dr Francesca Pizzo, Dr Sandrine Aubert, Dr Stanislas Lagarde, Dr Nada El Youssef, Dr Symela Chatzikonstantinou, Dr Francesca Bonini, Dr Angela Marchi, Dr Géraldine Daquin, Dr Lisa Vaugier, Dr Julia Makhalova, Dr Nathalie Villeneuve, Dr Anne Lepine, the medical residents and their teams for the clinical management of some of the included patients.

## Funding statement

The project leading to this publication has received support from the French government under the “France 2030” investment plan managed by the French National Research Agency (reference: ANR-16-CONV000X/ANR-17-EURE-0029) and from Excellence Initiative of Aix-Marseille University - A*MIDEX (AMX-19-IET-004).

## Conflict of interest statement

None of the authors has any conflict of interest to disclose.

## Ethics statement

All participants provided written informed consent prior to inclusion. The study was conducted in accordance with the Declaration of Helsinki and was approved by the local ethics committee of Assistance Publique–Hôpitaux de Marseille (AP-HM) (PADS25-96).

## Data availability

Data that support our findings are available from the corresponding author upon reasonable request.

## Notes

### Competing Interest Statement

The authors have declared no competing interest.

### Author Declarations

All participants provided written informed consent prior to inclusion. The study was conducted in accordance with the Declaration of Helsinki and was approved by the local ethics committee of Assistance Publique Hopitaux de Marseille (PADS25 96).

