## Supplemental Data 1 for "The burden of the postictal state in epilepsy: a prospective, single-centre observational cohort study"

**Supplementary materials - Table 1. Postictal Recovery Scale (PRS) (English translation):**

| **Domain** | **Question** | **Score 0** | **Score 1** | **Score 2** | **Score 3** |
| --- | --- | --- | --- | --- | --- |
| **1. Fatigue** | **To what extent do you feel tired after the seizure?** | Extremely tired; I cannot keep my eyes open or move | Very tired, with limited ability to move or concentrate | A little tired but able to move and respond | No unusual fatigue |
| **2. Emotional state: sadness/apathy** | **Do you feel sadness or apathy after the seizure?** | Very intense sadness or apathy; overwhelming and making it difficult to engage with the world | Noticeable sadness or apathy, but manageable | Mild sadness or apathy that I can manage without difficulty | No sadness or apathy |
| **3. Emotional state: anxiety** | **Do you feel anxiety after the seizure?** | Very intense anxiety; overwhelming and difficult to control | Noticeable anxiety, but manageable | Mild anxiety that I can manage without difficulty | No anxiety |
| **4. Emotional state: euphoria/ecstasy** | **Do you experience unusual happiness, euphoria or ecstasy after the seizure?** | Very strong ecstasy or euphoria, overwhelming and difficult to control, disturbing my concentration | Marked euphoria or strong elation, but still controllable | Slightly elevated and positive mood, above normal | No positive emotional change |
| **5. Sensory deficits** | **Do you notice changes in your senses (vision, hearing, touch, taste, smell) after the seizure?** | Severe sensory deficit (e.g. blindness, deafness, loss of sensation) | Major sensory deficit (e.g. very blurred vision, muffled hearing, altered touch) | Mild sensory disturbance (e.g. slight blurring or tingling) | No sensory deficit |
| **6. Motor and/or coordination deficits** | **To what extent can you move your body and perform actions after the seizure?** | Unable to move, or marked weakness in one or more limbs | Major weakness; movements are possible but difficult | Mild weakness; able to perform tasks but with difficulty | No motor deficit; my movements are normal and coordinated |
| **7. Spoken language comprehension** | **To what extent do you understand spoken language after the seizure?** | Unable to understand spoken language | Limited comprehension; I only understand very simple words or sentences | I understand most of it, but with some effort | Full comprehension, without difficulty |
| **8. Spoken language production** | **To what extent can you speak and communicate verbally after the seizure?** | Unable to speak or form words | I produce sounds or words, but have difficulty forming coherent sentences | I speak and form sentences, but with difficulty | No difficulty speaking |
| **9. Orientation in time and space** | **Do you feel oriented in time and space after the seizure?** | Very disoriented | I know where I am, but remain confused about time (or vice versa) | Oriented in time and space, with slight confusion | Perfectly oriented |
| **10. Time perception** | **How do you experience the passage of time after the seizure?** | No perception of the passage of time | Time seems very slow, as if minutes were stretching out | Time feels somewhat strange, but I can form an approximate sense of it | Time perception is normal; time passes as expected |
| **11. Postictal amnesia** | **Do you remember what happened during the seizure?** | I remember nothing of what happened during the seizure | I have vague and blurred memories of the events of the seizure | I remember some details, but there are gaps | I remember all details of the seizure |
| *Each of the 11 domains is scored from 0 to 3, with lower scores indicating greater postictal dysfunction. The total score ranges from 0 to 33.* | | | | | |

**Additional items (not included in score)**

| **Item** | **Open field** |
| --- | --- |
| **Sensory deficits** | *Please specify the affected senses and affected body side (± part(s)), if applicable:* __________ |
| **Motor and/or coordination deficits** | *Please specify the affected body side (± part(s)), if applicable:* __________ |
| **Other disturbances that you feel after the seizure?** | *Please specify if applicable:* __________ |

**Postictal recovery duration**
**When did you feel completely back to your normal (preictal) state?**

- 0–5 minutes after the seizure
- 5–10 minutes after the seizure
- 10–15 minutes after the seizure
- 15–30 minutes after the seizure
- 30–60 minutes after the seizure
- More than 1 hour after the seizure
- If known, please specify: __________
