## Supplemental Data 2 for "The burden of the postictal state in epilepsy: a prospective, single-centre observational cohort study"

| **Supplementary materials - Table 2. Classification of antiseizure medications according to their principal mechanism of action and distribution in the study cohort (n = 96).** | | | |
| --- | --- | --- | --- |
| **Anti-seizure medication** | **Main mechanism of action** | **Number of patients** | **Proportion of patients** |
| Lacosamide | Sodium-channel blocker | 42 | 44% |
| Cenobamate | Mixed | 34 | 35% |
| Lamotrigine | Sodium-channel blocker | 24 | 25% |
| Carbamazepine | Sodium-channel blocker | 21 | 22% |
| Levetiracetam | SV2A ligand | 20 | 21% |
| Brivaracetam | SV2A ligand | 19 | 20% |
| Clobazam | GABAergic | 19 | 20% |
| Eslicarbazepine | Sodium-channel blocker | 16 | 17% |
| Perampanel | Other | 10 | 10% |
| Topiramate |  | 9 | 9% |
| Valproate | Mixed | 8 | 8% |
| Zonisamide | Mixed | 5 | 5% |
| Oxcarbazepine | Sodium-channel blocker | 4 | 4% |
| Cannabidiol | Other | 3 | 3% |
| Clonazepam | GABAergic | 3 | 3% |
| Phenobarbital | GABAergic | 3 | 3% |
| Gabapentin | Other | 2 | 2% |
| Phenytoin | Sodium-channel blocker | 2 | 2% |
| Acetazolamide | Other | 1 | 1% |
| Diazepam | GABAergic | 1 | 1% |
